# Determining the Impact of Ethnicity on the Accuracy of Measurements of Oxygen Saturations. A Retrospective Cohort Study

**DOI:** 10.1101/2021.11.21.21266662

**Authors:** Mansoor N Bangash, James Hodson, Felicity Evison, Jaimin M Patel, Andrew McD Johnston, Suzy Gallier, Elizabeth Sapey, Dhruv Parekh

## Abstract

**Background:** Pulse oximeters are routinely used in community and hospital settings worldwide as a rapid, non-invasive, and readily available bedside tool to approximate blood oxygenation. Potential racial biases in SpO2 measurements may influence the accuracy of pulse oximetry readings and impact clinical decision making. We aimed to assess whether the accuracy of oxygen saturation measured by peripheral pulse oximetry (SpO2), relative to arterial blood gas (SaO2), varies by ethnicity.

**Methods:** In this large retrospective observational cohort study covering four NHS Hospitals serving a large urban population in Birmingham, consecutive patients admitted to hospital requiring oxygen therapy were identified by electronic patient records. For each spell, the first available pair of SpO2 and SaO2 measurements taken within an interval of less than 20 minutes were identified and included in the analysis. The differences between SpO2 and SaO2 measurements, were compared across self-identified groups of ethnicities. These differences were subsequently adjusted for age, sex, bilirubin, systolic blood pressure, carboxyhaemaglobin saturations and the time interval between SpO2 and SaO2 measurements.

**Findings:** Paired O2 saturation measurements from 16818 inpatient spells between 1st January 2017 and 18th February 2021, were analysed. The cohort self-identified as being of White (81.2%), Asian (11.7%), Black (4.0%), or Other (3.2%) ethnicities. Across the cohort, SpO2 was significantly higher than SaO2 (median: 98% vs. 97%, p<0.001), with a median difference of 0.5 percentage points (pps). However, the size of this difference varied considerably with the magnitude of SaO2, with SpO2 overestimation by a median of 3.8pp for SaO2 values <90% but underestimating by a median of 0.4pp for an SaO2 of 95%. The differences between SpO2 and SaO2 were also found to vary significantly by ethnicity, with this difference being 0.8pp (95% confidence interval: 0.6-1.0) greater in those of Black vs. White ethnicity. These differences resulted in 6.1% vs. 8.7% of White vs. Black patients classified as normoxic on SpO2 who were hypoxic on the gold standard SaO2 reading (p=0.007).

**Interpretation:** Pulse oximetry tends to overestimate O2 saturation, and this is more pronounced in patients of Black ethnicity. Prospective studies are urgently warranted to assess the impact of ethnicity on the accuracy of pulse oximetry, to ensure care is optimised for all.

**Funding:** This work was supported by PIONEER, the Health Data Research UK (HDR-UK) Health Data Research Hub in acute care. HDR-UK is an initiative funded by the UK Research and Innovation, Department of Health and Social Care (England) and the devolved administrations, and leading medical research charities.

## Introduction

Ensuring optimal oxygen delivery to tissues is a prime objective of acute/critical medical care. Using arterial blood gas (ABG) sampling and co-oximetry analysis to measure arterial blood oxygen saturation (SaO2) is considered the gold standard means of assessing blood oxygenation.^1^ However, ABG measurement is an invasive and labour-intensive test. Pulse oximeter measured oxygen saturation (SpO2) is a non-invasive approximation of SaO2. First developed in 1974^2^, pulse oximeters have been routinely used in both community and hospital settings for the past forty years, and are widely acknowledged to be highly important clinical monitoring devices that improve patient safety.^3^

The measurement of oxygen saturation by pulse oximetry is based upon two principles. The first is that oxyhaemoglobin (O2Hb) and deoxyhaemoglobin (Hb) have different absorption spectra, with O2Hb absorbing greater amounts of infrared light and lower amounts of red light, compared to Hb. The second is that beat-to-beat variation of tissue blood volume produces a light transmission signal which depends only on the pulse characteristics of arterial blood. Pulse oximeters emit two wavelengths of light, red at 660 nm and near-Infrared at 940 nm, from a pair of light-emitting diodes located in one side of the probe, which is usually placed on a finger (over the nailbed). The transmission of the two wavelengths of light through the finger is then detected by a photodiode on the opposite arm of the probe, and the relative amount of red and infrared light absorbed are ultimately used to determine the proportion of Hb bound to oxygen.^4^ Pulse oximetry only detects the oxygen saturation of arterial blood, through subtraction of non-varying, venous (deoxygenated) blood spectra, due to the recognition of fluctuations in absorbed light caused by the cardiac cycle.^5^

The accuracy of pulse oximetry readings is vital. Overestimation of actual SaO2 might lead to clinically relevant hypoxaemia remaining undetected and untreated. Conversely, underestimation of actual SaO2 may result in unnecessary oxygen therapy, with the potential risks of hyperoxaemia^6^, oxygen wastage and inappropriate clinical decision making, including severity of illness assessment and safe discharge from hospital. The United States Regulatory body which governs approvals for medical devices, the Food and Drug Administration (FDA), requires the accuracy of pulse oximeters to be tested against SaO2, with the root-mean-square difference between the SpO2 and the true SaO2 being within ±2-3%.^7^ There are known conditions where SpO2 readings may not accurately reflect the true SaO2. These include, but are not limited to, methaemoglobinaemia^8^, severe carbon monoxide poisoning^9^, hyperbilirubinaemia^5^, excessive movement, hypotension and hypoperfusion states, severe anaemia, and increased blood glycohaemoglobin.^5 10 11^

A recent study has highlighted potential racial biases in SpO2 measurements.^12^ In this study, the frequency of occult hypoxemia that was not detected by SpO2 was almost three times higher in Black patients, compared to White patients. There have been previous, much smaller studies assessing the impact of skin pigmentation on the accuracy of SpO2 compared to SaO2, but often with discordant results.^13-15^

Given the widespread use of SpO2 in clinical decision making worldwide, guidance suggesting narrow therapeutic windows for oxygen therapy^16^, and care pathways placing more emphasis on early discharge, ambulatory and home monitoring for patient management, any systemic differences between SpO2 and SaO2 could have major health implications. On the other hand, adjustments to measurements on the basis of ethnicity (for example, the estimated glomerular filtration) are now increasingly controversial^17 18^, and there has been considerable interest in revalidating scoring systems to remove references to ethnicity.^19^ As such, any changes to clinical practice or test interpretation driven by ethnicity requires a sound evidence base.

Birmingham, UK is one of the most diverse urban centres in the UK.^20^ The aim of this study was to assess whether SpO2 accurately predicted SaO2, measured by ABG, and whether disparities between ethnicities existed.

## Methods

### Study design and population

This data study was supported by PIONEER, a Health Data Research Hub in Acute Care with ethical approval provided by the East Midlands – Derby REC (reference: 20/EM/0158).

University Hospitals Birmingham NHS Foundation Trust (UHB), UK, is one of the largest hospital complexes in Europe, covering four NHS hospital sites, treating over 2.2 million patients per year, and housing the largest single critical care unit (CCU) in Europe. UHB provides secondary care to a diverse population of 1.3 million in Birmingham and Solihull, and provides a full range of tertiary services to the West Midlands region. UHB runs a fully electronic healthcare record (EHR) (PICS; Birmingham Systems), which has been in place since 1999. Data within PICS is time- and date-stamped to the nearest millisecond, and includes all physiology and arterial blood gas analysis outputs.

### Case selection and data collection

Between 1^st^ Jan 2017 and 18^th^ February 2021, data were collected for all measurements of oxygen saturation by oximetry and ABG made during inpatient spells at UHB. From these, pairs of measurements (SpO2 and SaO2) on the same patient taken within an interval of less than 20 minutes were identified. All consecutive measurements meeting these criteria were initially included in the study, to reduce the risk of selection bias as much as possible. In order to minimise the effect of within-spell correlation of outcomes, particularly for those patients with extended lengths of stay, only a single pair of measurements per spell were included in the analysis, namely the first valid pair of measurements after admission. Any cases with an O2 saturation by either measure of <80% were subsequently excluded, as these were deemed likely to be spurious results (for example, misreported venous blood gas measurements for SaO2).

Patient demographics and clinical data were collected from the EHR and from mandatory data sets within the Hospital Trust. Ethnicity was self-reported by the patient or their family members on admission to hospital.

### O2 Saturation measurements

ABG analysers automatically update the EHR, with SaO2 measurements being recorded to one decimal place. No such linkage existed for pulse oximeters; hence SpO2 measurements were collected rounded to the nearest integer, and manually entered into the EHR by health care professionals. For the analyses of differences between the two approaches, calculations were made to one decimal place. However, for analysis of the absolute differences between approaches, the SaO2 values were rounded to the nearest integer, prior to calculations being performed. In addition, the exclusion criteria of O2 saturation <80% used a value of <79.5% for SaO2, for consistency with SpO2.

### Statistical methods

Initially, measurements on SpO2 and SaO2 were compared using Wilcoxon’s signed rank test, with the strength of the correlation quantified using Spearman’s (rho) coefficient. Since O2 saturations are recorded as percentages, differences between SpO2 and SaO2 were reported as percentage point (pp) differences. These were calculated as SpO2 minus SaO2, such that positive differences represented higher values on SpO2. To test for any proportional bias, the differences between SpO2 and SaO2 were also quantified within subgroups of SaO2 measurements. To model this relationship, a generalized additive model (GAM) was produced, with the pp difference between measures as the dependent variable, and the smooth function of the SaO2 as a continuous covariate. A second model was also produced with the SpO2 as the dependent variable. Since this followed a negatively skewed distribution, SpO2 values were subtracted from 101 to reverse the skew, before being log_2_-transformed, in order to normalise the distribution, and ensure reliability of the model.

Cohort characteristics and O2 saturations were then compared across groups of ethnicity using Kruskal-Wallis tests for continuous variables and Chi-square tests for nominal variables. The association between ethnicity and the differences between SpO2 and SaO2 were then further interrogated. Since the differences between SpO2 and SaO2 varied by the magnitude of the measurement, the previously described GAM model was extended to additionally include ethnicity as a nominal factor. This model was then further extended to adjust for the effects of other potentially confounding factors. The goodness of fit was assessed graphically for each factor, with transformations (e.g. log_2_) applied, where poor fit was identified.

Due to the complexities of modelling the relationship between SpO2 and SaO2, an alternative approach was also used as a sensitivity analysis. Here, SaO2 values were dichotomised, into hypoxic (SaO2<94.0%) and normoxic (SaO2≥94.0%), with a threshold of 94.0% being selected since this is a commonly used clinical oxygenation threshold in acutely unwell patients without chronic respiratory disease.^21^ This was then set as the dependent variable in a binary logistic regression model, with SpO2 as a continuous covariate, and ethnicity as a nominal factor. To further assess the ability for SpO2 to identify hypoxia, the proportions of patients misclassified by SpO2, relative to SaO2, were calculated for each ethnicity, and compared using chi-square tests.

Continuous variables are reported as (arithmetic) mean ± standard deviation (SD) where approximately normally distributed, or as median (interquartile range; IQR) otherwise. Analyses were performed using IBM SPSS 24 (IBM Corp. Armonk, NY), with GAM modelling performed using the R package “mgcv”. Cases with missing data were excluded from analyses of the affected variable, and p<0.05 was deemed to be indicative of statistical significance throughout.

### Sample size calculation

After exclusions, data were available for N=16818 cases, with the differences between SpO2 and SaO2 having a standard deviation of 3.1pp (see ***Results*** section for further details). Based on these values, the study was sufficiently powered to detect a difference in the mean of SpO2 minus SaO2 between patients of White (81.2% of cohort) and Black (4.0%) ethnicity of 0.4pp, at 80% power and 5% alpha.

### Patient and public involvement

This project was discussed by a multi-ethnic group of patients and members of the public, including the background and rationale for the project, the proposed analysis on the basis of self-defined ethnic groups and the results to date. There was overwhelming support for this work including the use of potentially sensitive data on ethnicity, given the clinical importance and public interest in the topic. This PPIE group will also help write lay summaries of the results to increase public awareness of the outputs.

## Results

### Cohort characteristics

Pairs of SpO2 and SaO2 measurements were available for a total of N=20231 inpatient spells in N=18069 patients. Of these, the ethnicity was not recorded in N=2612 (12.9%) spells, and these were excluded from further analysis. Of the remaining N=17619 cases, N=689 (3.9%) had an SaO2<80%, and N=178 (1.0%) had an SpO2<80%; N=66 (0.4%) of these had O2 saturations <80% on both measures. The distributions of O2 saturations <80% were similar across ethnicities (p=0.077, ***Supplementary Table 1***). After excluding these cases, a total of N=16,818 pairs of O2 saturation measurements were included in subsequent analysis (**Figure 1**).

**Figure 1.**
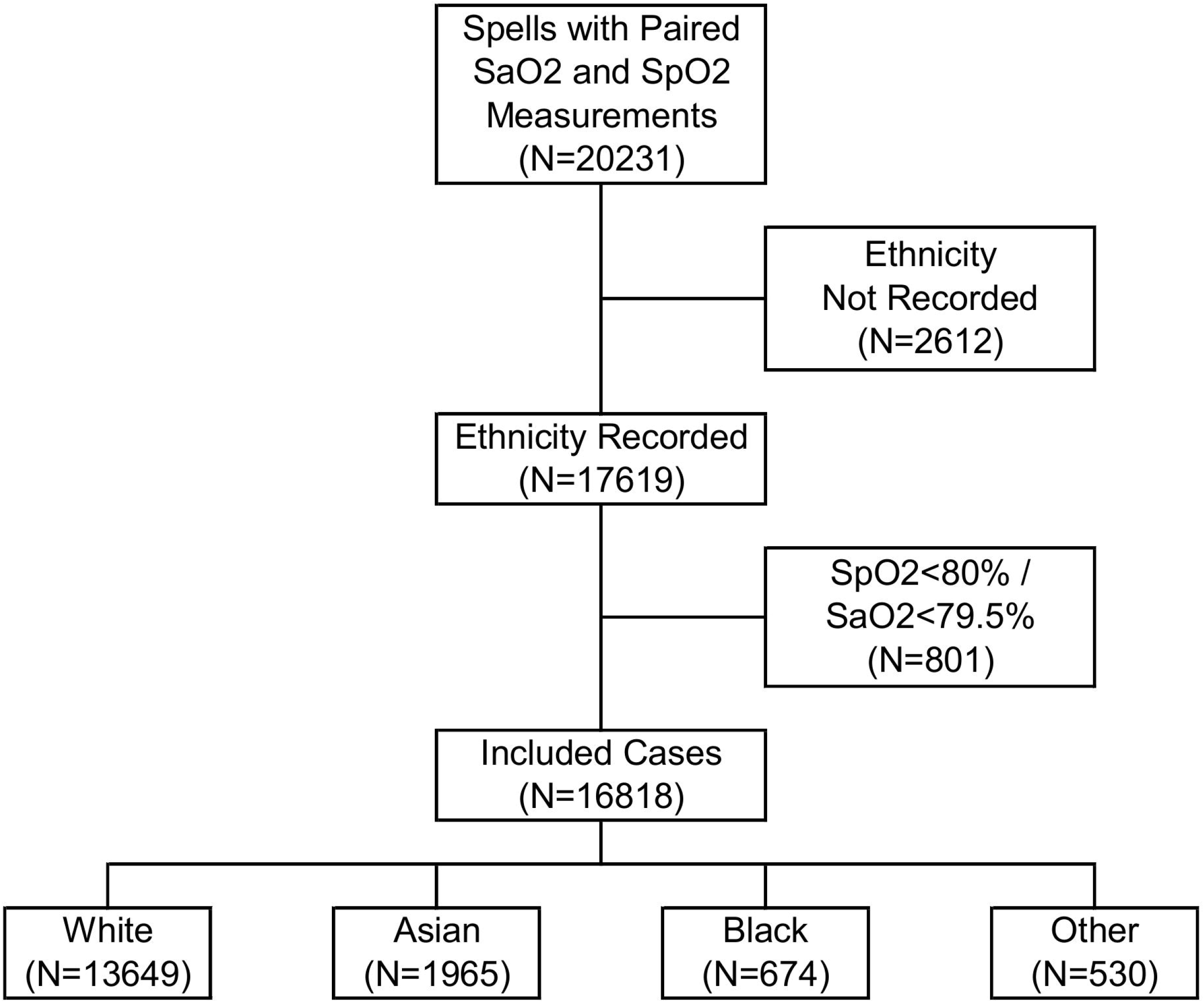
Study flowchart.

The median patient age was 63 years (IQR: 50-74), and 57.9% of cases were male. The majority of patients were of self-reported White ethnicity (N=13649; 81.2%), with the remainder self-reporting Asian (N=1965; 11.7%), Black (N=674; 4.0%) or other (mixed non-White: N=530; 3.2%) ethnicities. See ***Table 1*** for cohort demography and physiological data.

**Table 1.** Cohort characteristics by ethnicity.

|  | N | Total Cohort | Ethnicity |  |  |  | p-Value |
| --- | --- | --- | --- | --- | --- | --- | --- |
|  |  |  | White | Asian | Black | Other |  |
| Age (Years) | 16818 | 63 (50, 74) | 65 (53, 75) | 57 (42, 68) | 55 (45, 68) | 50 (35, 64) | **<0.001** |
| < 50 |  | 4101 (24.4%) | 2878 (21.1%) | 729 (37.1%) | 231 (34.3%) | 263 (49.6%) |  |
| 50 - 64 |  | 4700 (27.9%) | 3740 (27.4%) | 578 (29.4%) | 243 (36.1%) | 139 (26.2%) |  |
| 65 - 74 |  | 4045 (24.1%) | 3556 (26.1%) | 348 (17.7%) | 67 (9.9%) | 74 (14.0%) |  |
| 75 - 84 |  | 2866 (17.0%) | 2506 (18.4%) | 235 (12.0%) | 85 (12.6%) | 40 (7.5%) |  |
| 85+ |  | 1106 (6.6%) | 969 (7.1%) | 75 (3.8%) | 48 (7.1%) | 14 (2.6%) |  |
| Sex (% Male)* | 16815 | 9735 (57.9%) | 7796 (57.1%) | 1217 (61.9%) | 388 (57.6%) | 334 (63.0%) | **<0.001** |
| FiO2 (fraction of inspired air) | 16733 | 0.28 (0.21, 0.40) | 0.28 (0.21, 0.40) | 0.28 (0.21, 0.45) | 0.28 (0.21, 0.40) | 0.28 (0.21, 0.45) | **<0.001** |
| PaO2 (kPa)** | 16543 | 12.7 (10.0, 17.6) | 12.6 (10.0, 17.4) | 13.4 (10.2, 19.0) | 12.9 (10.2, 18.8) | 13.0 (10.1, 18.3) | **<0.001** |
| Bilirubin (μmol/L) | 16488 | 10 (7, 17) | 10 (7, 18) | 9 (6, 15) | 9 (6, 15) | 10 (7, 17) | **<0.001** |
| Systolic BP (mmHg) | 15948 | 124 ± 26 | 124 ± 26 | 124 ± 26 | 132 ± 29 | 125 ± 26 | **<0.001** |
| MetHb (%) | 16680 | 0.6 (0.5, 0.7) | 0.6 (0.5, 0.7) | 0.6 (0.5, 0.7) | 0.6 (0.5, 0.7) | 0.6 (0.5, 0.7) | 0.631 |
| COHb (%) | 16681 | 1.4 (1.1, 1.7) | 1.4 (1.1, 1.7) | 1.2 (0.9, 1.5) | 1.3 (1.1, 1.7) | 1.3 (1.1, 1.6) | **<0.001** |
| Time SpO2 to SaO2 (Minutes) |  |  |  |  |  |  |  |
| SaO2 Minus SpO2 | 16818 | 0.0 (-7.4, 7.8) | 0.0 (-7.5, 7.8) | 0.0 (-6.9, 8.0) | 0.0 (-7.0, 8.1) | 0.0 (-8.0, 7.3) | 0.639 |
| Absolute Difference | 16818 | 7.6 (2.9, 13.0) | 7.6 (2.9, 13.0) | 7.5 (2.6, 12.9) | 7.6 (3.0, 12.8) | 7.5 (3.2, 13.1) | 0.615 |
| O2 Saturation (%) |  |  |  |  |  |  |  |
| SaO2 | 16818 | 97.4 (95.6, 98.7) | 97.4 (95.6, 98.6) | 97.6 (95.6, 98.8) | 97.5 (95.7, 98.8) | 97.6 (95.6, 98.8) | **0.001** |
| SpO2 | 16818 | 98 (95, 100) | 97 (95, 100) | 98 (95, 100) | 99 (96, 100) | 98 (95, 100) | **<0.001** |
| Difference in O2 Saturation (pp) |  |  |  |  |  |  |  |
| SpO2 Minus SaO2 | 16818 | 0.5 (-1.2, 1.4) | 0.4 (-1.3, 1.4) | 0.7 (-0.7, 1.5) | 0.8 (-0.3, 1.9) | 0.7 (-1.0, 1.6) | **<0.001** |
| Absolute Difference*** | 16818 | 1 (1, 3) | 1 (1, 3) | 1 (1, 2) | 1 (1, 3) | 1 (1, 3) | 0.348 |

Continuous variables are reported as median (interquartile range) or mean ± standard deviation, with p-values from Kruskal-Wallis tests. Nominal variables are reported as N (column %), with p-values from Chi-square tests. Bold p-values are significant at $p < 0.05$ . \*vs. female – excludes N=3 who identified as Non-Binary. \*\*Temperature-corrected. \*\*\* SaO2 values were rounded to the nearest integer when calculating absolute differences, for consistency with the SpO2 measurements. pp=percentage points; SaO2=arterial O2 saturation; SpO2=O2 saturation on oximetry. FiO2=fraction of inspired oxygen; PaO2=partial pressure of oxygen; BP=blood pressure; MetHb=methaemoglobin; COHb=carboxyhaemoglobin.

### O2 saturations on SpO2 vs. SaO2

O2 saturations were found to follow a highly negatively skewed distribution; however, the shape of this distribution varied between measurement methods (***Figure 2a***). On SpO2, 30.2% (N=5087) of cases had an O2 saturation of 100%, compared to only 5.4% (N=915) of cases on SaO2 (i.e. SaO2≥99.5%). The median SpO2 was 98% (IQR: 95-100%), which was significantly higher than the 97.4% (IQR: 95.6-98.7%) on SaO2 (p<0.001). The correlation between SpO2 and SaO2 was relatively modest, with Spearman’s rho of 0.663.

**Figure 2.**
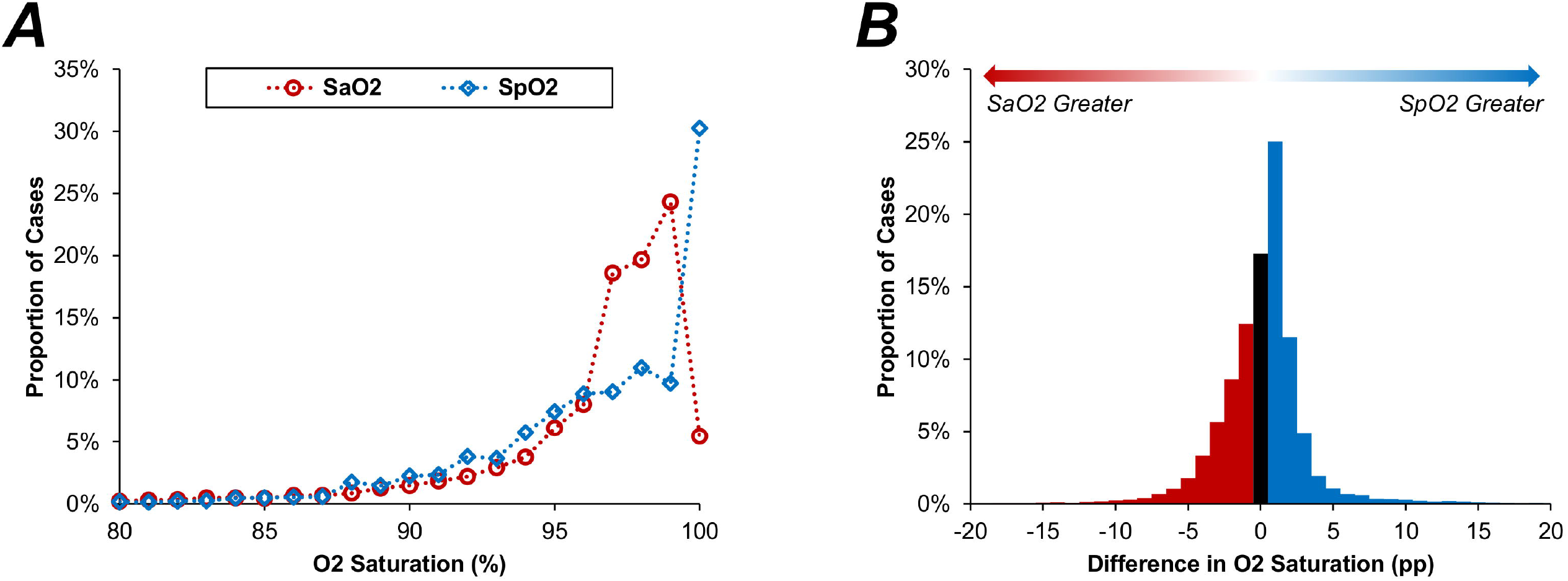
Distributions of SaO2 and SpO2. In Figure A, points represent the proportion of cases with each integer value of O2 saturation; SaO2 values were rounded to the nearest integer before plotting. Figure B is a histogram of the percentage point (pp) differences in SpO2 minus SaO2 (with SaO2 values rounded to the nearest integer). Each bar represents 1 pp of O2 saturation, with the black bar representing concordance between SpO2 and SaO2 (i.e. a difference of zero). SaO2=arterial O2 saturation; SpO2=O2 saturation on oximetry

The differences in O2 saturations between measurement methods (SpO2 minus SaO2) were assessed in further detail. These differences were approximately normally distributed, albeit with long tails, with a median difference of 0.5pp (IQR: -1.2, 1.4) and a mean difference of 0.1 ± 3.1pp (***Figure 2b***). After rounding the SaO2 values to the nearest integer, O2 saturations were concordant on the two measurement approaches in only 17.3% (N=2910) of cases, with SpO2 giving the greater value in 47.6% (N=8,008), and SaO2 being greater in 35.1% (N=5,900).

The relationship between SpO2 and SaO2 was then assessed within subgroups of SaO2, in order to test for any proportional bias. This found the difference between measurement methods to vary with the magnitude, following a complex trend, which is visualised in ***Table 2*** and ***Figure 3***. For the lowest values of SaO2 (i.e. <89.5%), SpO2 tended to be greater than SaO2, with a median difference of 3.8pp (IQR: 0.4, 8.8). The size of this difference then reduced with increasing SaO2, with no significant difference between median measurements on SpO2 vs. SaO2 where SaO2 values were in the range 91.5-92.4% (p=0.639) or 92.5-93.4% (p=0.876). However, SpO2 subsequently began to underestimate, relative to SaO2, with a median difference of -0.4pp (IQR: -2.0, 1.4; p<0.001) for the subgroup with SaO2 of 94.5-95.4%. The direction of this effect then reversed again, with SpO2 tending to give higher values than SaO2 for the subgroup with SaO2 of 97.5-98.4% (median difference: 0.6pp; IQR: -0.9, 1.8).

**Table 2.** Associations between SaO2 and SpO2.

| SaO2 (%) | N<br>Pairs | Difference in O2 Saturation<br>(SpO2 minus SaO2; pp) |  | O2<br>Saturation<br>Higher on*: | p-Value |
| --- | --- | --- | --- | --- | --- |
| | | Mean ± SD | Median (IQR) | | |
| < 89.5 | 952 | 4.7 ± 5.6 | 3.8 (0.4, 8.8) | SpO2 | **<0.001** |
| 89.5 - 90.4 | 247 | 1.4 ± 4.0 | 1.1 (-1.4, 3.9) | SpO2 | **<0.001** |
| 90.5 - 91.4 | 308 | 0.8 ± 3.8 | 0.6 (-1.8, 3.1) | SpO2 | **0.002** |
| 91.5 - 92.4 | 369 | 0.2 ± 3.5 | -0.2 (-2.2, 2.3) | NS | 0.639 |
| 92.5 - 93.4 | 487 | 0.0 ± 3.3 | -0.2 (-2.0, 1.9) | NS | 0.876 |
| 93.5 - 94.4 | 631 | -0.4 ± 3.2 | -0.3 (-2.2, 1.7) | SaO2 | **0.013** |
| 94.5 - 95.4 | 1029 | -0.4 ± 3.1 | -0.4 (-2.0, 1.4) | SaO2 | **<0.001** |
| 95.5 - 96.4 | 1351 | -0.4 ± 2.8 | -0.2 (-1.8, 1.6) | SaO2 | **<0.001** |
| 96.5 - 97.4 | 3128 | -0.3 ± 2.5 | -0.1 (-1.6, 1.5) | SaO2 | **0.021** |
| 97.5 - 98.4 | 3311 | 0.1 ± 2.2 | 0.6 (-0.9, 1.8) | SpO2 | **<0.001** |
| 98.5 - 99.4 | 4090 | -0.1 ± 2.2 | 0.7 (-0.5, 1.1) | SpO2 | **<0.001** |
| $\geq 99.5$ | 915 | -0.8 ± 2.7 | 0.3 (-0.6, 0.5) | NS | 0.456 |

Subgroup analyses were performed within intervals of SaO2. For each subgroup, the percentage point (pp) differences between SpO2 and SaO2 are reported as both mean ± standard deviation (SD), and as median (interquartile range; IQR), and compared using Wilcoxon's test. Bold p-values are significant at $p < 0.05$ . \*The measure with the higher O2 saturation, based on the sums of positive vs. negative ranks from the Wilcoxon's test. NS = no significant difference; SaO2=arterial O2 saturation; SpO2=O2 saturation on oximetry; pp=percentage points

**Figure 3.**
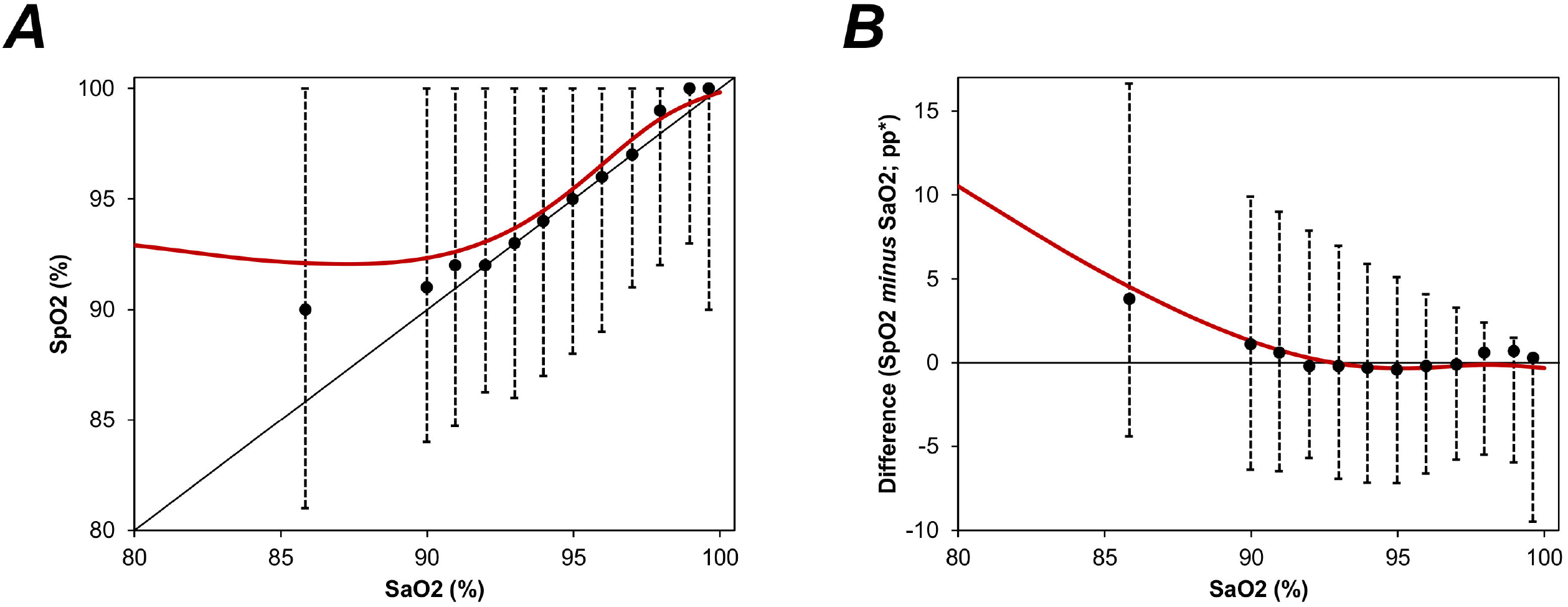
Association between SaO2 and SpO2. Points represent the median SpO2 within intervals of SaO2. The first point includes SaO2 measurements <89.5%, with subsequent intervals having a width of 1 percentage point (89.5-90.4%, 90.5-91.4% etc.). Whiskers represent the 2.5 th and 97.5 th percentiles, hence the range comprising 95% of values. Red lines are from generalized additive models (GAMs), with the smooth function of SaO2 as the independent variable, and either log2(101-SpO2) or the percentage point difference between SpO2 and SaO2 as the dependent variable. *The O2 saturation on SpO2 minus SaO2, as a percentage point (pp) difference. SaO2=arterial O2 saturation; SpO2=O2 saturation on oximetry.

This subgroup analysis also showed discrepancies between the average difference when quantified as a mean or median. For example, within the subgroup of patients with a SaO2 of 98.5-99.4%, the median difference was positive (0.7pp), implying measurements were higher on SpO2, whilst the mean difference was negative (−0.1pp), implying measurements were lower on SpO2. This occurred since, whilst the differences between methods was approximately normally distributed for the cohort as a whole (***Figure 2b***), the distribution became negatively skewed for high O2 saturations, largely since there was an upper bound at an O2 saturation of 100%. This is visualised in the ridgeline plot in ***Figure 4***.

**Figure 4.**
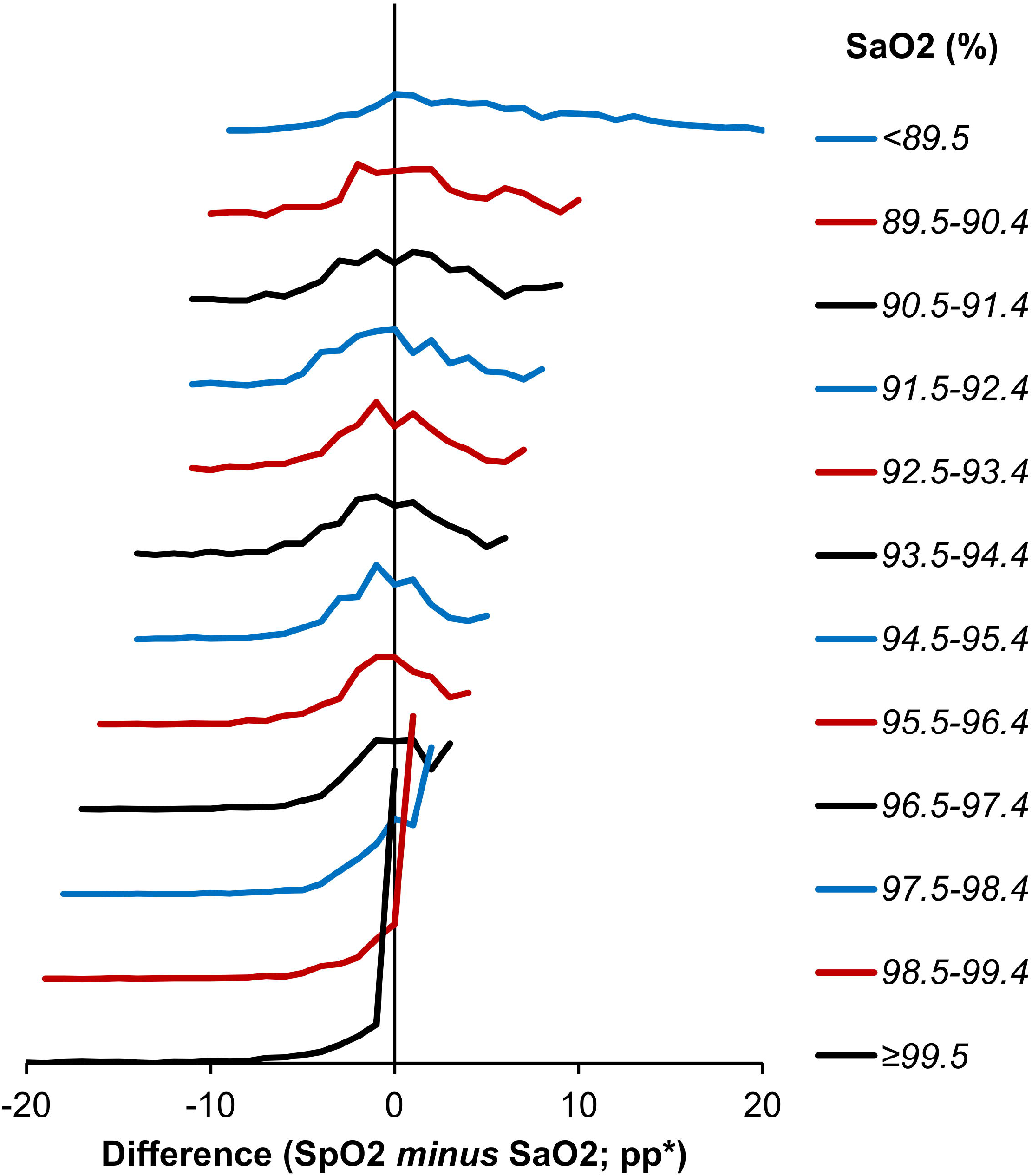
Ridgeline plot of the differences in O2 saturations by SaO2. Lines represent the distributions of the differences in O2 saturations within subgroups of SaO2, and are truncated at the minimum and maximum observed values. *Differences in O2 saturations are calculated as SpO2 minus SaO2, and are reported as percentage point (pp) differences. SaO2=arterial O2 saturation; SpO2=O2 saturation on oximetry.

### Cohort characteristics by ethnicity

Comparisons of patient characteristics across the groups of ethnicity found a significant difference in age (p<0.001), with a median of 65 years in White patients, compared to medians of 50-57 years in the non-White ethnic groups (p<0.001, ***Table 1***). A significant difference in the sex distribution was also observed (p<0.001), with a greater preponderance of males in the Asian and “other” ethnic groups. Bilirubin was found to be significantly higher in White patients (p<0.001), whilst those of Black ethnicity had significantly higher systolic BP (p<0.001). The time intervals between SpO2 and SaO2 measurements were similar across the ethnicities, with the median absolute differences ranging from 7.5 to 7.6 minutes (p=0.615).

### Differences in O2 saturations by ethnicity

Whilst SaO2 was found to differ significantly between groups (p=0.001), the magnitude of this difference was relatively small, with medians ranging from 97.4% in White patients to 97.6% in the Asian group. However, the difference in SpO2 was more pronounced, with medians ranging from 97% in White patients, to 99% in Black patients (p<0.001). As such, the differences between SpO2 and SaO2 varied significantly with ethnicity, with SpO2 being a median of 0.4pp higher than SaO2 in White patients, compared to 0.8pp higher in those of Black ethnicity (p<0.001, ***Table 1***).

Since the difference between SpO2 and SaO2 had previously been shown to vary by the magnitude of the measurement, analyses were then performed to compare this between ethnicities, after adjusting for magnitude of SaO2. The resulting model (***Figure 5a***) found the significant difference between ethnicities to persist, after adjusting for the magnitude of the measurement (p<0.001). Relative to patients of White ethnicity, the differences between SpO2 and SaO2 were 0.5pp (95% CI: 0.4-0.7, p<0.001) greater in Asian patients, 0.8pp (0.6-1.0, p<0.001) greater in Black patients, and 0.3pp (0.1-0.6, p=0.006) greater in those other ethnicities.

**Figure 5.**
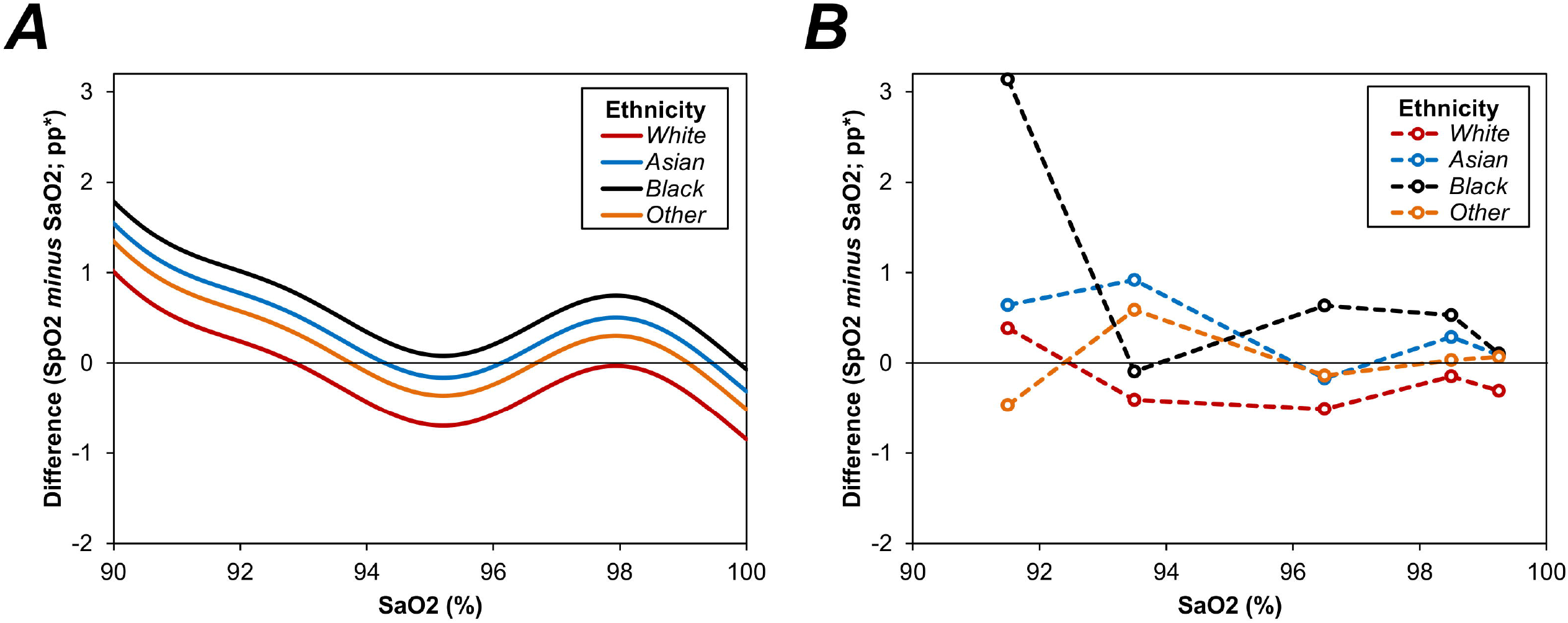
Association between SaO2 and SpO2 by ethnicity. Figure A represents a generalized additive model (GAM), with the percentage point (pp) difference in O2 saturations between SpO2 and SaO2 as the dependent variable, the smooth function of SaO2 as a covariate, and ethnicity as a nominal factor. Figure B visualises the observed mean differences between SpO2 and SaO2 within intervals of SaO2, to validate the goodness of fit of the GAM model. Each interval had a width of 2pp, and points are plotted at the midpoint of the interval. Both plots are left-truncated at an SaO2 of 90%, in order to better visualise the differences between groups. *Differences in O2 saturations are calculated asSpO2 minus SaO2, and are reported as percentage point (pp) differences. SaO2=arterial O2 saturation; SpO2=O2 saturation on oximetry.

This model was then further extended, to adjust for other potentially confounding factors (***Table 3***). This found the difference in O2 saturations (SpO2 minus SaO2) to be significantly more positive in males (p<0.001), younger patients (p<0.001), those with lower COHb (p<0.001), and those with higher systolic BP (p=0.041). After adjusting for these factors, the differences between ethnicities remained significant (p<0.001), with more positive differences between measures of O2 saturations observed for Asian (0.3pp, p<0.001) and Black (0.6pp, p<0.001) patients, relative to those of White ethnicity.

**Table 3.** Associations with percentage point differences in O2 saturation between SpO2 and SaO2.

|  | Analysis 1) Univariable |  | Analysis 2) SaO2 + Ethnicity |  | Analysis 3) All Factors |  |
| --- | --- | --- | --- | --- | --- | --- |
|  | Coefficient (95% CI) | p-Value | Coefficient (95% CI) | p-Value | Coefficient (95% CI) | p-Value |
| SaO2 | N/A* | <0.001 | N/A* | <0.001 | N/A* | <0.001 |
| Ethnicity |  | <0.001 |  | <0.001 |  | <0.001 |
| White | - | - | - | - | - | - |
| Asian | 0.49 (0.34, 0.63) | <0.001 | 0.53 (0.40, 0.67) | <0.001 | 0.33 (0.19, 0.47) | <0.001 |
| Black | 0.83 (0.59, 1.07) | <0.001 | 0.78 (0.56, 0.99) | <0.001 | 0.62 (0.40, 0.84) | <0.001 |
| Other | 0.33 (0.07, 0.60) | 0.015 | 0.33 (0.09, 0.57) | 0.006 | 0.11 (-0.14, 0.35) | 0.394 |
| Age (per Decade) | -0.12 (-0.15, -0.09) | <0.001 |  |  | -0.20 (-0.22, -0.17) | <0.001 |
| Sex (Male) | 0.03 (-0.06, 0.13) | 0.509 |  |  | 0.15 (0.07, 0.24) | <0.001 |
| Bilirubin (per Two-Fold Increase)** | -0.05 (-0.08, -0.01) | 0.015 |  |  | 0.01 (-0.03, 0.04) | 0.630 |
| Systolic BP (per 10 mmHg) | 0.03 (0.01, 0.05) | <0.001 |  |  | 0.02 (0.00, 0.03) | 0.041 |
| COHb (per 10pp) | -1.14 (-1.70, -0.59) | <0.001 |  |  | -3.11 (-3.64, -2.59) | <0.001 |
| Time from SpO2 to SaO2 (per 10 mins) | -0.03 (-0.08, 0.01) | 0.172 |  |  | -0.01 (-0.05, 0.04) | 0.745 |

### Associations between SpO2 and normoxia by ethnicity

Due to the complexities of modelling the relationship between SpO2 and SaO2, the analysis of ethnicity was repeated using an alternative approach as a sensitivity analysis. Initially, the SaO2 values were dichotomised using a cut-off value of 94.0%, with the N=14,165 (84.2%) of cases that were greater than or equal to this threshold being classified as normoxic, and the remainder as hypoxic. A binary logistic regression model was then produced, to assess the relationship between SpO2 and normoxia on SaO2 (***Figure 6a***). This model was then extended to include ethnicity as a factor (***Figure 6b, Table 4, Table 5***), which was found to be statistically significant (p<0.001). For a given SpO2 value, both Asian (odds ratio [OR]: 0.75, p<0.001) and Black (OR: 0.67, p=0.003) patients were significantly less likely to be normoxic, compared to those of White ethnicity, implying a greater tendency for SpO2 to overestimate O2 saturations in these non-White groups

**Table 4.**
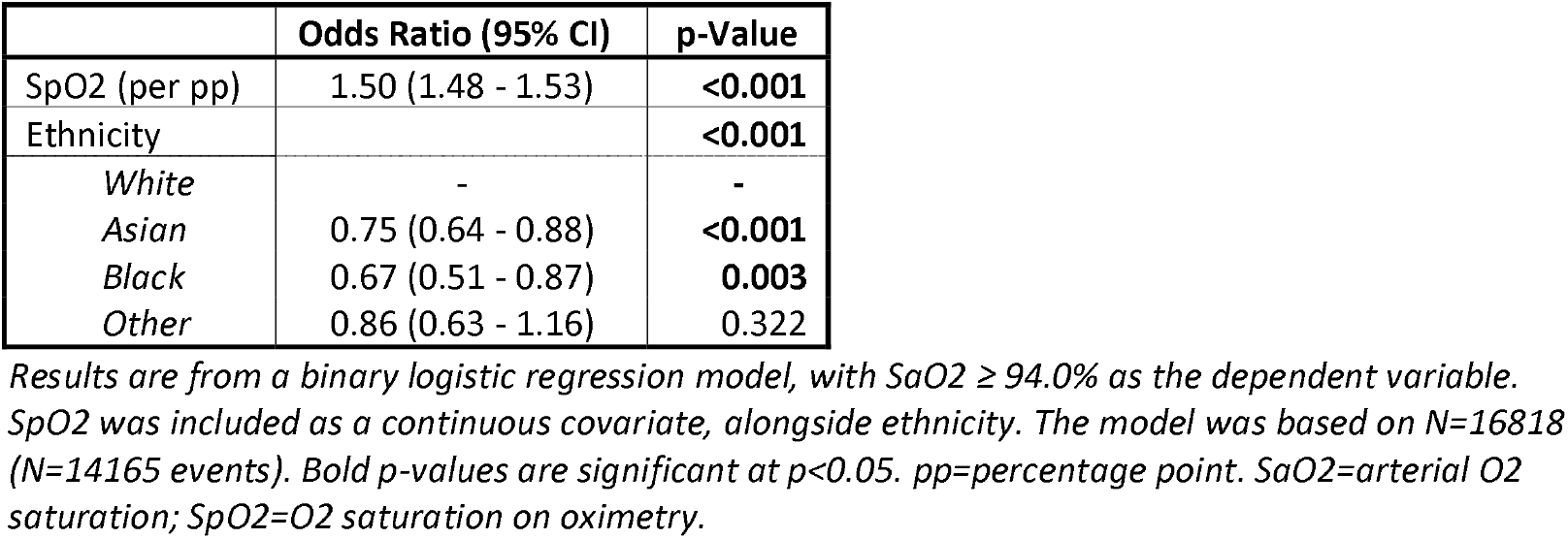
Binary logistic regression model of normoxia (SaO2 ≥ 94.0%)

|  | Odds Ratio (95% CI) | p-Value |
| --- | --- | --- |
| SpO2 (per pp) | 1.50 (1.48 - 1.53) | **<0.001** |
| Ethnicity |  | **<0.001** |
| White | - | - |
| Asian | 0.75 (0.64 - 0.88) | **<0.001** |
| Black | 0.67 (0.51 - 0.87) | **0.003** |
| Other | 0.86 (0.63 - 1.16) | 0.322 |

Results are from a binary logistic regression model, with SaO2 ≥ 94.0% as the dependent variable. SpO2 was included as a continuous covariate, alongside ethnicity. The model was based on N=16818 (N=14165 events). Bold p-values are significant at p<0.05. pp=percentage point. SaO2=arterial O2 saturation; SpO2=O2 saturation on oximetry.

**Table 5.** Association between SpO2 and rates of normoxia (SaO2 ≥ 94.0%) by ethnicity.

| SpO2 | Cases With SaO2 ≥ 94.0% |  |  |  |
| --- | --- | --- | --- | --- |
|  | White | Asian | Black | Other |
| <90% | 23.4% (197/843) | 29.6% (24/81) | 24.2% (8/33) | 25.0% (10/40) |
| 90% | 31.8% (100/314) | 31.3% (10/32) | 29.4% (5/17) | 54.5% (6/11) |
| 91% | 41.1% (139/338) | 21.4% (9/42) | 70.0% (7/10) | 50.0% (2/4) |
| 92% | 53.2% (280/526) | 49.4% (40/81) | 40.0% (6/15) | 50.0% (7/14) |
| 93% | 65.4% (334/511) | 63.8% (44/69) | 68.8% (11/16) | 75.0% (12/16) |
| 94% | 78.8% (651/826) | 76.6% (72/94) | 75.0% (18/24) | 78.3% (18/23) |
| 95% | 86.9% (936/1077) | 80.0% (88/110) | 72.7% (24/33) | 69.7% (23/33) |
| 96% | 91.0% (1140/1253) | 86.7% (124/143) | 89.4% (42/47) | 86.0% (43/50) |
| 97% | 94.7% (1183/1249) | 87.6% (156/178) | 88.5% (46/52) | 90.5% (38/42) |
| 98% | 95.7% (1474/1541) | 94.9% (186/196) | 92.3% (60/65) | 93.5% (43/46) |
| 99% | 97.1% (1288/1327) | 95.8% (183/191) | 93.7% (74/79) | 94.9% (37/39) |
| 100% | 97.9% (3765/3844) | 97.2% (727/748) | 94.7% (268/283) | 97.6% (207/212) |

Data were initially divided into subgroups, based on the value of SpO2. Within each subgroup, the % (n/N) of patients with SaO2 ≥ 94.0% is then reported for each ethnicity. SaO2=arterial O2 saturation; SpO2=O2 saturation on oximetry.

**Figure 6.**
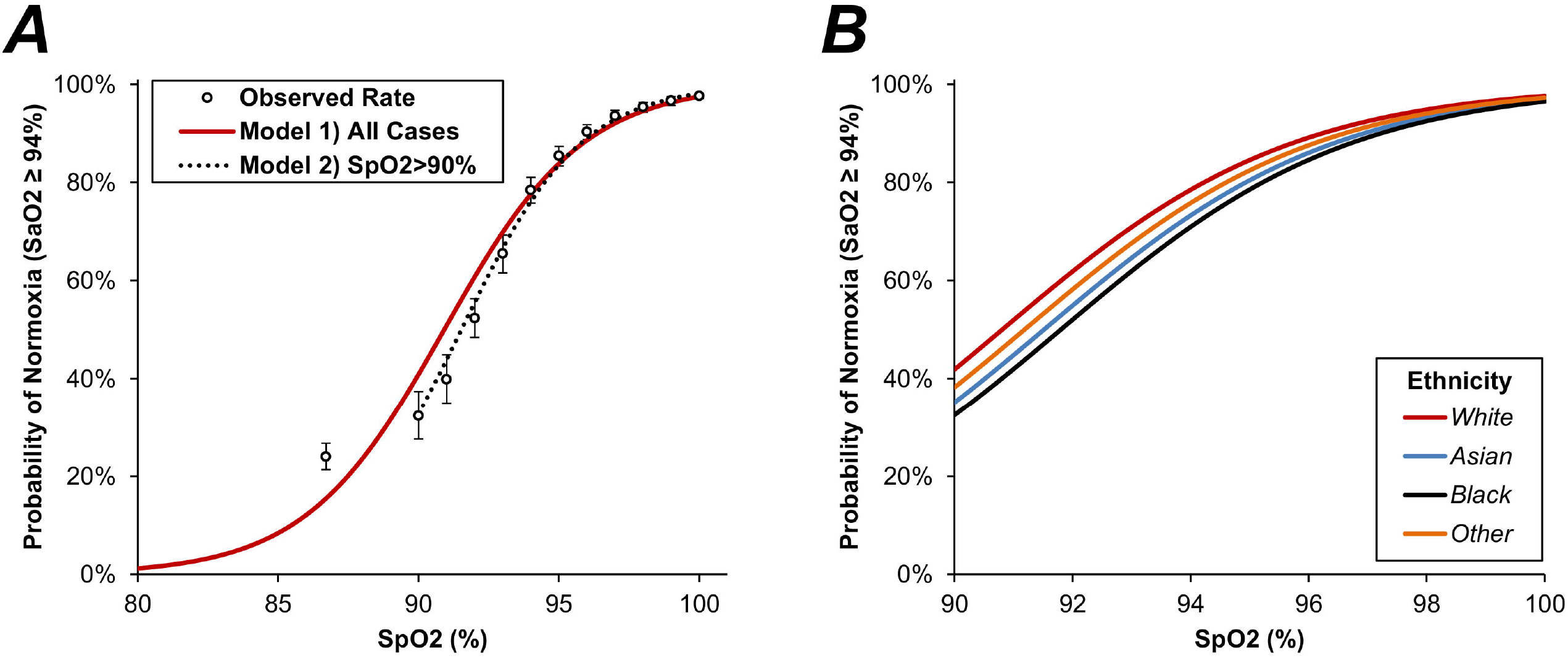
Association between SpO2 and the probability of normoxia (SaO2 ≥ 94.0%) In Figure A, points represent the probability of SaO2 ≥ 94.0% for SpO2 of 80-89% (first point), and for each integer value of SpO2 from 90-100%. Whiskers represent 95% confidence intervals. The red line is from a binary logistic regression model, with SpO2 as a continuous covariate. This model appeared to have suboptimal fit in for low values of SpO2, due to the greater inaccuracy of oximetry in this range. As such, a second model (broken line) was produced which only included the subset with SpO2>90%, which returned similar results in this range. In Figure B, the first binary logistic regression model was extended to include ethnicity as a factor, see **Table 4** for further details; this figure is truncated at an SpO2 of 90%, to more clearly highlight the difference between groups. SaO2=arterial O2 saturation; SpO2=O2 saturation on oximetry.

In order to further quantify this effect, a misclassification analysis was performed (***Table 6***). For the cohort as a whole, 41.5% (1251/3013) of those with SpO2<94% had been misclassified, and were actually normoxic on SaO2; this rate was similar across the subgroups of ethnicities (p=0.982). In those with SpO2 ≥ 94%, a total of 6.5% (891/13805) had been misclassified, and were hypoxic on SaO2. This rate was found to differ significantly with ethnicity (p=0.007), with SpO2 misclassifying 6.1% (680/11117) of White patients as normoxic, compared to 8.7% (51/583) of Black patients.

**Table 6.** Misclassification analysis.

| Subgroup | SpO2 | Hypoxia<br>(SaO2 < 94.0%) | Normoxia<br>(SaO2 ≥ 94.0%) |
| --- | --- | --- | --- |
| **Overall** | <94% | 1762 (58.5%) | 1251 (41.5%) |
|  | ≥94% | 891 (6.5%) | 12914 (93.5%) |
| **By Ethnicity** |  |  |  |
| **White** | <94% | 1482 (58.5%) | 1050 (41.5%) |
|  | ≥94% | 680 (6.1%) | 10437 (93.9%) |
| **Asian** | <94% | 178 (58.4%) | 127 (41.6%) |
|  | ≥94% | 124 (7.5%) | 1536 (92.5%) |
| **Black** | <94% | 54 (59.3%) | 37 (40.7%) |
|  | ≥94% | 51 (8.7%) | 532 (91.3%) |
| **Other** | <94% | 48 (56.5%) | 37 (43.5%) |
|  | ≥94% | 36 (8.1%) | 409 (91.9%) |

Cases were initially classified as hypoxic or normoxic, based on SaO2. The distribution of hypoxia vs. normoxia was then calculated within those cases with SpO2 <94% and ≥94% for the cohort as a whole, as well as within each subgroup of ethnicity. Data are reported as N (row %).

## Discussion

This study reports paired oxygen saturation measurements from pulse oximetry and ABG (the accepted gold standard) from almost 17000 inpatient spells at a secondary care NHS trust with four hospitals, which serves a large diverse urban catchment area. This is the largest study conducted to assess this issue, to date.

The relationship between SpO2 and SaO2 measurements was complex, and varied across the O2 saturation spectrum (80% to 100%). At lower values of O2 saturation (SaO2 80% - 91.4%), SpO2 considerably overestimated oxygenation (relative to SaO2). At higher values (SaO2 93.5-97.4%), SpO2 considerably underestimated O2 saturation. These differences were exaggerated in measurements taken from Black ethnic groups, and misclassification (classified as normoxic when hypoxic) occurred significantly more often in this group compared to those of White ethnicity. Similar (albeit smaller) effects were observed in those of Asian ethnicity compared to White patients. Despite the complex relationship between SpO2 and SaO2, these differences between ethnicities appeared to persist over the range of O2 saturations.

The discrepancies at low oxygen saturations are likely to be clinically significant and the impact could be important. An SaO2 reading of 86% was often associated with a falsely reassuring SpO2 of >90%, leaving hypoxaemia potentially undetected and untreated. Analysis of misclassification types suggests that SpO2 measurements that impact a reduction in oxygen therapy are less affected across ethnicities than scenarios where oxygen therapy may need to be applied or escalated.

Our results are consistent with a much smaller study of 11 healthy people with darkly pigmented skin and 10 healthy people with light skin pigmentation, who were oxygen restricted as part of a physiological study. This study tested three different pulse oximeters, all of which gave higher readings, relative to SaO2, in those with dark- vs. light-pigmented skin.^22^ A larger follow-on study of 36 participant replicated these findings and, importantly, highlighted differences in performance of devices and variations amongst differing levels of skin pigmentation (dark, light and intermediate). This is replicated in our findings of Asian participants having less discrepancy than Black participants. Our results are also consistent with results from a large study reported in 2020 ^12^. They reported paired measures of oxygen saturation by pulse oximetry and ABG obtained from two cohorts: one comprising 1333 White patients and 276 Black patients from 2020, and one multi-centre cohort comprising 7342 White patients and 1050 Black patients from 2014 – 2015, finding a consistent difference of 2% across the spectrum of SpO2 measurements in Black compared to White patients. Our study not only includes patients of all ethnicities, but also contains a more detailed statistical analysis of the relationship between SpO2 and SaO2 at different clinically relevant cut-off values of SpO2.

Strengths of our study include its size and representation of a multi-cultural, urban population in the UK across 4 hospital sites. The primary limitation of this study was its retrospective nature, with the baseline cohort comprising patients who had undergone assessment of O2 saturation by both oximetry and ABG, which may not be representative of the inpatient cohort as a whole. In addition, initial assessment of the data identified O2 saturations that were unfeasibly low, and likely to represent either erroneous or incorrectly recorded measurements. In an attempt to exclude these spurious values, a minimum O2 saturation of 80% on both oximetry and ABG was selected as a threshold, with measurements below this being excluded. However, if this approach is too strict, then genuine measurements may have been excluded, introducing selection bias, whereas if it was too lenient, then erroneous data may have been included in the analysis.

A second limitation of the study is the potential for confounding, due to the differences in baseline characteristics between the ethnicity groups, for example, the large difference in average age. This age discrepancy has been described in other studies of hospitalised patients where ethnicity was a focus^23^ and, therefore, may reflect true differences in admitted populations, but this could impact on the results of the current study. Whilst multivariable analyses were employed to adjust for these differences, modelling will always leave some degree of residual confounding. There is also the potential for other intangible or unmeasured confounding factors to have influenced the findings of the analysis. Thirdly, the SpO2 and SaO2 readings were separated by on average 7.5 minutes and were not all simultaneous readings. There is a degree of SpO2 dynamic variation, especially in acutely ill patients that this study design could not account for. Fourthly, the self-reported groups of ethnicity used in the analysis were reasonably broad. As such, whilst these may act as a surrogate of skin colour, there is likely to be considerably within-group variability. If the observed discrepancies in oximetry measurements, relative to SaO2 are, to some degree, a consequence of the degree of skin pigmentation, then this may have acted as a confounding factor in the analysis. Finally, the data included in the study comprised inpatient spells from hospitals based in the same geographical area and operated under the same NHS trust. As such, the results may not be generalizable to other hospitals, particularly if there are large differences in patient demographics.

Health inequalities due to ethnicity have been highlighted during the COVID-19 pandemic^23^. There is an increasing drive for early discharges from hospital, as well as for remote home monitoring to avoid unnecessary admissions^24 25^. Pulse oximetry is used routinely in all healthcare settings, and is the mainstay of rapid quantification of a patient’s oxygenation status. Although O2 saturations measured on pulse oximetry should never be used in isolation for clinical decision-making, they are included in many severity scoring systems and decision-making algorithms, such as those for pneumonia and COPD. Our findings highlight the need for some caution, as there is potential risk of overestimation of SpO2 measurement, which appears to be particularly pronounced in patients in ethnicities with a tendency for darker skin pigmentation. This could potentially lead to inadequate treatment if, for example, insufficient oxygenation in patients COVID-19 is missed, leading to patients being sent home, rather than admitted, potentially delaying or denying access to evidence-based therapies.

## Conclusions

This study highlights differences in the measurement of oxygenation status when using pulse oximeters and arterial blood gases. Discrepancies are largest when oxygen saturations are lowest, where SpO2 consistently overestimates SaO2. Across the range of O2 saturations, the difference between SpO2 and SaO2 is exaggerated in those of Black ethnicity, potentially placing them at most risk of undetected hypoxic events. Whilst there is insufficient evidence to change current practice, caution may need to be exercised in some small, specific subgroups of patients with borderline SpO2 levels. Prospective studies are urgently warranted to further quantify the degree of any such racial bias in current devices, to ensure care is optimised for all.

## Research in Context

### Evidence before this study

We searched PubMed for studies of ethnic disparities in oxygen saturations, published up to Sept 31, 2021. We used the search terms ([“Ethnicity” or “Racial”] and [“Oximetry” OR “Saturations” OR “Oxygen”]). We found one large study from US centres reporting paired measures of oxygen saturation by pulse oximetry and ABG obtained from two cohorts: one comprising 1333 White patients and 276 Black patients and one multi-centre cohort comprising 7342 White patients and 1050 Black patients from 2014 – 2015, finding a consistent difference of 2% across the spectrum of SpO2 measurements in Black compared to White patients. Other studies have been small and in defined disease states. Disparities in all ethnic groups and differences at clinically relevant cut-off values of SpO2 remain unconfirmed yet important due to the widespread use of pulse oximetry in clinical decision making for admission and discharge to hospital.

### Added Value of this Study

This is the largest cohort study in a diverse cohort in terms of ethnicity, over a significant period and in a spectrum of acute illness severity and admission reason. Providing real-life and internationally translatable data. We highlight differences in the measurement of oxygenation status when using pulse oximeters and arterial blood gases. Discrepancies are largest when oxygen saturations are lowest, where SpO2 consistently overestimates SaO2. Across the range of O2 saturations, the difference between SpO2 and SaO2 is exaggerated in those of Black ethnicity, potentially placing them at most risk of undetected hypoxic events. Similar but smaller effects were observed in those of Asian ethnicity compared to White patients. Despite the complex relationship between SpO2 and SaO2, these differences between ethnicities appeared to persist over the range of O2 saturations.

### Implications of all the available evidence

Our findings and others suggest that validation of pulse oximeters should focus on non-White racial populations, in order to identify any shortcomings and drive technological refinements. Whilst newer and more accurate devices are being developed, we urge companies and regulatory authorities to provide performance characteristics across ethnicities, and quantify any bias that may be present, so that clinicians can be fully informed in decision-making.

### Ethical Statement

#### Ethical Approval

This data study was supported by PIONEER, a Health Data Research Hub in Acute Care with ethical approval provided by the East Midlands – Derby REC (reference: 20/EM/0158).

### Data Availability Statement

Data is available upon reasonable application to the PIONEER Hub. All data access applications will be reviewed in accordance with the PIONEER ethical approvals and governance processes.

## Supporting information

STROBE Checklist

## Contributors

DP, MB, and ES were responsible for the study design. JH, SG, FC did the data analyses. DP, ES, and JH drafted the manuscript, and all authors contributed to critical revision of the manuscript for important intellectual content and approved it for submission. DP and ES are the guarantors. The corresponding author attests that all listed authors meet authorship criteria and that no others meeting the criteria have been omitted.

## Competing interests

All authors have completed the ICMJE uniform disclosure form at www.icmje.org/coi_disclosure.pdf and declare: DP reports funding from the NIHR and MRC. ES reports funding support from HDR-UK, MRC, Wellcome Trust, NIHR, Alpha-1-Foundation and British Lung Foundation. ES declares receiving consulting fees from Boehringer Ingleheim and support to attend scientific meetings from Astra Zeneca. MB reports funding from the UK Intensive Care Society

The lead author affirms that this manuscript is an honest, accurate, and transparent account of the study being reported; that no important aspects of the study have been omitted; and that any discrepancies from the study as planned (and, if relevant, registered) have been explained.

**Supplementary Table 1.** Exclusions by ethnicity.

| O2 Saturation |  | Ethnicity |  |  |  |
| --- | --- | --- | --- | --- | --- |
| *SpO2* | *SaO2* * | *White*<br>(*N=14300*) | *Asian*<br>(*N=2049*) | *Black*<br>(*N=723*) | *Other*<br>(*N=547*) |
| ≥80% | ≥79.5% | 13649 (95.4%) | 1965 (95.9%) | 674 (93.2%) | 530 (96.9%) |
| <80% | ≥79.5% | 92 (0.6%) | 12 (0.6%) | 6 (0.8%) | 2 (0.4%) |
| ≥80% | <79.5% | 501 (3.5%) | 66 (3.2%) | 41 (5.7%) | 15 (2.7%) |
| <80% | <79.5% | 58 (0.4%) | 6 (0.3%) | 2 (0.3%) | 0 (0.0%) |

*Chi-square test: p=0.077. \*A cut-off value of 79.5%, rather than 80% was used for ABG, since values were reported to one decimal place. SaO2=arterial O2 saturation; SpO2=O2 saturation on oximetry.*

## Notes

### Author Declarations

This data study was supported by PIONEER, a Health Data Research Hub in Acute Care with ethical approval provided by the East Midlands Derby REC (reference: 20/EM/0158).

## References

1. Pilcher J, Ploen L, McKinstry S, et al. A multicentre prospective observational study comparing arterial blood gas values to those obtained by pulse oximeters used in adult patients attending Australian and New Zealand hospitals. BMC Pulm Med 2020;20(1):7. doi: 10.1186/s12890-019-1007-3 [published Online First: 2020/01/11]

2. Aoyagi T, Miyasaka K. Pulse oximetry: its invention, contribution to medicine, and future tasks. Anesth Analg 2002;94(1 Suppl):S1–3. [published Online First: 2002/03/20]

3. Jubran A. Pulse oximetry. Crit Care 1999;3(2):R11–R17. doi: 10.1186/cc341 [published Online First: 2000/11/30]

4. Sinex JE. Pulse oximetry: principles and limitations. Am J Emerg Med 1999;17(1):59–67. doi: 10.1016/s0735-6757(99)90019-0 [published Online First: 1999/02/03]

5. Chan ED, Chan MM, Chan MM. Pulse oximetry: understanding its basic principles facilitates appreciation of its limitations. Respir Med 2013;107(6):789–99. doi: 10.1016/j.rmed.2013.02.004 [published Online First: 2013/03/16]

6. Palmer E, Post B, Klapaukh R, et al. The Association between Supraphysiologic Arterial Oxygen Levels and Mortality in Critically Ill Patients. A Multicenter Observational Cohort Study. Am J Respir Crit Care Med 2019;200(11):1373–80. doi: 10.1164/rccm.201904-0849OC [published Online First: 2019/09/13]

7. Food and Drug Administration. Pulse Oximeters – Premarket Notification Submissions: Guidance for Industry and Food and Drug Administration Staff. https://www.fdagov/downloads/MedicalDevices/DeviceRegulationandGuidance/GuidanceDocuments/UCM081352pdf2013; Accessed August 2021

8. Barker SJ, Tremper KK, Hyatt J. Effects of methemoglobinemia on pulse oximetry and mixed venous oximetry. Anesthesiology 1989;70(1):112–7. doi: 10.1097/00000542-198901000-00021 [published Online First: 1989/01/01]

9. Hampson NB. Pulse oximetry in severe carbon monoxide poisoning. Chest 1998;114(4):1036–41. doi: 10.1378/chest.114.4.1036 [published Online First: 1998/10/29]

10. Hinkelbein J, Koehler H, Genzwuerker HV, et al. Artificial acrylic finger nails may alter pulse oximetry measurement. Resuscitation 2007;74(1):75–82. doi: 10.1016/j.resuscitation.2006.11.018 [published Online First: 2007/03/14]

11. Pu LJ, Shen Y, Lu L, et al. Increased blood glycohemoglobin A1c levels lead to overestimation of arterial oxygen saturation by pulse oximetry in patients with type 2 diabetes. Cardiovasc Diabetol 2012;11:110. doi: 10.1186/1475-2840-11-110 [published Online First: 2012/09/19]

12. Sjoding MW, Dickson RP, Iwashyna TJ, et al. Racial Bias in Pulse Oximetry Measurement. N Engl J Med 2020;383(25):2477–78. doi: 10.1056/NEJMc2029240 [published Online First: 2020/12/17]

13. Adler JN, Hughes LA, Vivilecchia R, et al. Effect of skin pigmentation on pulse oximetry accuracy in the emergency department. Acad Emerg Med 1998;5(10):965–70. doi: 10.1111/j.1553-2712.1998.tb02772.x [published Online First: 1998/12/23]

14. Foglia EE, Whyte RK, Chaudhary A, et al. The Effect of Skin Pigmentation on the Accuracy of Pulse Oximetry in Infants with Hypoxemia. J Pediatr 2017;182:375–77 e2. doi: 10.1016/j.jpeds.2016.11.043 [published Online First: 2016/12/13]

15. Emery JR. Skin pigmentation as an influence on the accuracy of pulse oximetry. J Perinatol 1987;7(4):329–30. [published Online First: 1987/01/01]

16. Siemieniuk RAC, Chu DK, Kim LH, et al. Oxygen therapy for acutely ill medical patients: a clinical practice guideline. BMJ 2018;363:k4169. doi: 10.1136/bmj.k4169 [published Online First: 20181024]

17. Diao JA, Inker LA, Levey AS, et al. In Search of a Better Equation - Performance and Equity in Estimates of Kidney Function. N Engl J Med 2021;384(5):396–99. doi: 10.1056/NEJMp2028243 [published Online First: 2021/01/07]

18. Eneanya ND, Yang W, Reese PP. Reconsidering the Consequences of Using Race to Estimate Kidney Function. JAMA 2019;322(2):113–14. doi: 10.1001/jama.2019.5774 [published Online First: 2019/06/07]

19. Diao JA, Wu GJ, Taylor HA, et al. Clinical Implications of Removing Race From Estimates of Kidney Function. JAMA 2021;325(2):184–86. doi: 10.1001/jama.2020.22124 [published Online First: 2020/12/03]

20. Birmingham City Council. Population and Census. https://www.birminghamgovuk/info/20057/about_birmingham/1294/population _and_census 2011;Accessed August 2021

21. O’Driscoll BR, Howard LS, Earis J, et al. BTS guideline for oxygen use in adults in healthcare and emergency settings. Thorax 2017;72(Suppl 1):ii1–ii90. doi: 10.1136/thoraxjnl-2016-209729

22. Bickler PE, Feiner JR, Severinghaus JW. Effects of skin pigmentation on pulse oximeter accuracy at low saturation. Anesthesiology 2005;102(4):715–9. doi: 10.1097/00000542-200504000-00004 [published Online First: 2005/03/26]

23. Sapey E, Gallier S, Mainey C, et al. Ethnicity and risk of death in patients hospitalised for COVID-19 infection in the UK: an observational cohort study in an urban catchment area. BMJ Open Respir Res 2020;7(1) doi: 10.1136/bmjresp-2020-000644 [published Online First: 2020/09/03]

24. NHSx. Supporting care with remote monitoring. https://www.nhsxnhsuk/covid-19-response/technology-nhs/supporting-the-innovation-collaboratives-to-expand-their-remote-monitoring-plans/ 2021;DOA 17th August 2021

25. Chapman ALN, Patel S, Horner C, et al. Updated good practice recommendations for outpatient parenteral antimicrobial therapy (OPAT) in adults and children in the UK. JAC Antimicrob Resist 2019;1(2):dlz026. doi: 10.1093/jacamr/dlz026 [published Online First: 2019/08/26]

